# Both general- and central- obesity are causally associated with polycystic ovarian syndrome: Findings of a Mendelian randomization study

**DOI:** 10.1101/2022.02.07.22270650

**Authors:** Kushan De Silva, Ryan T. Demmer, Daniel Jönsson, Aya Mousa, Helena Teede, Andrew Forbes, Joanne Enticott

**Author notes:** **Name and contact information of the corresponding author** Kushan De Silva, **Postal Address**, Locked Bag 29, Level 1, 43-51 Kanooka Grove, Clayton VIC 3168, Australia, Monash Centre for Health Research and Implementation, School of Public Health and Preventive Medicine, Faculty of Medicine, Nursing and Health Sciences, Monash University, Australia, **Email:**.

## Abstract

**Introduction:** Obesity is observed in a majority of women with polycystic ovarian syndrome (PCOS). Using body mass index (BMI) as a proxy, previous Mendelian randomization studies revealed general obesity potentially causes PCOS. Central obesity frequently demonstrates a stronger association with PCOS, although evidence on its causality is sparse.

**Objectives:** To investigate causal effects of both central- and general- obesity on the development of PCOS via two-sample Mendelian randomization (2SMR).

**Methods:** Summary GWAS data of female-only, large-sample cohorts of European ancestry were retrieved for anthropometric markers of central obesity (waist circumference (WC), hip circumference (HC), waist-to-hip ratio (WHR)) and general obesity (BMI and its constituent variables – weight and height), from the IEU Open GWAS Project. As the outcome data, we acquired summary data from a large-sample GWAS (96391 samples; 219 cases and 96172 controls) from the FinnGen cohort. Four 2SMR methods were applied: inverse variance weighted (IVW); MR Egger (MRE); weighted median (WME); weighted mode (WMO). Single SNP-, leave-one-out-, heterogeneity-, horizontal pleiotropy- and outlier- analyses were conducted. Genetic architectures underlying causal associations were explored.

**Results:** All SNPs selected as instrumental variables demonstrated no weak instrument bias (F > 10). Three anthropometric exposures, namely, BMI (OR: 5.55 – 7.24, WC (OR: 6.79 – 24.56), and HC (OR: 6.78 – 24.56), significantly causally associated with PCOS as per IVW, WME, and WMO models. Single SNP- and leave-one-out- sensitivity analysis results were indicative of robust causal estimates. No significant heterogeneity, horizontal pleiotropy, and outliers were observed. We observed a considerable degree of overlap (7 SNPs; 17 genes) across significant causal findings as well as a number of SNPs and genes that were not shared between causal associations.

**Conclusions:** This study revealed that both and general- and central obesity potentially cause PCOS. Findings underscore the importance of addressing obesity and adiposity for the prevention and management of PCOS.

## 1. INTRODUCTION

Polycystic ovarian syndrome (PCOS) is the most common endocrine disorder in women of reproductive age [1], with a prevalence ranging 8%-18% among this group [2]. It is also the major cause of anovulatory female infertility [3]. While a multitude of reproductive and metabolic abnormalities associate with PCOS, its convoluted etiology, presumably multifactorial and heterogeneous, is still not completely known. Classic clinico-pathologic features of PCOS comprise hyperandrogenism, oligo-anovulation, excessive weight or obesity and a range of metabolic manifestations such as glucose intolerance, insulin resistance, and dyslipidemia while a number of obstetric, cardiometabolic, oncological, and psychological complications may also ensue [4]. A recent review highlighted the correlation between obesity, hyperandrogenism, and insulin resistance, and suggested these might act in concert forming a vicious cycle to induce PCOS [5]. In addition, neuroendocrine changes such as gonadotropin secretory abnormalities [6], fetal programming alterations within the intrauterine microenvironment [7], genetic- and epigenetic factors [8], an array of environmental predictors [9] and certain inflammatory regulators [10] have also been implicated in the pathogenesis of PCOS. Constraints to the present PCOS care practices include a lack of uniform diagnostic criteria, limitations intrinsic to the evidence base emanating from observational epidemiological studies such as the concerns of confounding and reverse causality [11] as well as diagnostic delays and difficulties [12].

It should be noted that a substantial proportion of the PCOS-related evidence base stems from observational studies which are precluded by various biases and unmeasured confounding. However, causal inference is fundamental to unravelling disease etiologies and mechanistic underpinnings. The gold standard of etiological inference remains randomized controlled trials which could overcome key limitations of observational studies. Given the practical drawbacks of randomized trials especially exorbitant financial costs and longer time spans entailed, Mendelian randomization (MR) methods offer a viable and robust alternative to inferring causality using observational data. Using genetic variants as instrumental variables (IVs) and leveraging on their strengths, especially stability and random assortment of alleles, the causality between risk factors and diseases can be plausibly deduced by MR. With the increasing availability of genome-wide association studies (GWAS) data, MR is making considerable contributions to uncovering complex, polygenic disease etiologies [13]. Well-conducted MR studies meeting the three key assumptions of relevance, independence, and exclusion restriction could thus provide high-level evidence on causality [14]. To this end, guidelines such as the Strengthening the Reporting of Observational Studies in Epidemiology Using Mendelian Randomization (STROBE-MR) [15] have been developed to facilitate uniform and comprehensive presentation of MR studies.

Obesity is frequently observed in women with PCOS, predominantly in the form of abdominal obesity, aggravating metabolic and reproductive sequalae and PCOS-associated complications [16]. Available evidence suggests that the close link between obesity and PCOS is presumably mediated by multiple mechanisms such as insulin resistance-driven metabolic changes, steroidogenic and reproductive effects of hyperinsulinemia, and augmented adipokine synthesis by subcutaneous and visceral fat [17]. De Segher et al underscored the absence of a gonadotropic and/or ovarian disorder in a majority of girls with PCOS and theorized that central obesity and central adiposity are the main drivers of PCOS development [18]. Putative genetic links between obesity and adiposity with PCOS are reinforced by current evidence [19-21], for instance, the influence of *FTO* gene variants [22-23]. A caveat in the relationship between obesity and PCOS is that obesity alone may be neither a necessary nor a sufficient condition for its development among all PCOS-susceptible females [19], given the presence of lean PCOS phenotypes [24], exemplifying the complex, multifactorial etiology of PCOS.

Notably, multiple MR studies revealed that obesity potentially causes PCOS, all of which exclusively used body mass index (BMI) as the surrogate anthropometric measure of obesity [25-28]. However, BMI is essentially a marker of general obesity whereas central obesity in particular seems to confer a formidable influence on PCOS pathogenesis [16-18]. Moreover, the use of multiple anthropometric traits could yield not only complementary but also exclusive information. For example, a recent consensus statement highlighted the unequivocal evidence that waist circumference (WC) provides additive and independent information to BMI and recommended that WC be used as a vital sign in clinical practice [29]. Also, concordant and consistent findings emanating from MR analyses using multiple anthropometric markers of central- and general-obesity could strengthen the evidence pertaining to their putative causal roles in ensuing PCOS.

In this study, we examined existing data from GWAS, and aimed to investigate the causal effects of both central- and general-obesity on the development of PCOS via a two-sample Mendelian randomization (2SMR) approach. Specifically, we used anthropometric markers of central obesity (WC, hip circumference (HC), waist-to-hip ratio (WHR)) and general obesity (BMI and its constituent variables – weight and height), as exposures in 2SMR analyses.

## 2. MATERIALS AND METHODS

This study was conducted according to STROBE-MR guidelines [15], as described in **S1 Table**.

### 2.1. Data sources for exposures

For the six anthropometric traits selected as exposures, we performed a comprehensive search on the IEU Open GWAS Project database (https://gwas.mrcieu.ac.uk/) to find the most suitable GWAS summary data for each 2SMR analysis. Given the female-exclusivity of PCOS and sex-based variations in genetically-proxied anthropometric traits [30], we retrieved female-only GWASs. In order to alleviate confounding by ancestry and population stratification effects, we resorted to cohorts with participants of European ancestry. We prioritized GWASs with large sample sizes and ultimately identified appropriate studies from the **G**enetic **I**nvestigation of **AN**thropometric **T**raits (GIANT) consortium (**Table 1**).

**Table 1:** Female-only, European ancestry GWASs retrieved from the IEU Open GWAS Project to be included as exposures (anthropometric markers) and the outcome (PCOS) in two-sample Mendelian randomization analyses.

| Exposures and outcome | GWAS ID | Consortium | Sample size | No: of SNPs | Reference |
| --- | --- | --- | --- | --- | --- |
| **Exposures: Anthropometric markers** |  |  |  |  |  |
| Weight (kg) | ieu-a-107 | GIANT | 73137 | 2747007 | Randall et al., 2013; PMID = 23754948 |
| Height (m) | ieu-a-97 | GIANT | 73137 | 2748546 | Randall et al., 2013; PMID = 23754948 |
| Body mass index (BMI)<br>(kg/m<sup>2</sup>) | ieu-a-974 | GIANT | 171977 | 2494613 | Locke et al., 2015; PMID = 25673413 |
| Waist circumference (WC)<br>(cm) | ieu-a-63 | GIANT | 127997 | 2444355 | Shungin et al., 2015; PMID = 25673412 |
| Hip circumference (HC)<br>(cm) | ieu-a-51 | GIANT | 127997 | 2444355 | Shungin et al., 2015; PMID = 25673412 |
| Waist-to-hip ratio (WHR) | ieu-a-75 | GIANT | 118003 | 2466102 | Shungin et al., 2015; PMID = 25673412 |
| **Outcome** |  |  |  |  |  |
| Polycystic ovarian syndrome (PCOS) | finn-a-E4_POCS | FINNGEN | 96391(219 cases; 96172 controls) | 16152119 | N/A |

For weight, we selected the study with the GWAS-ID “ieu-a-107” reporting summary statistics from a meta-analysis of 46 cohorts over 73137 females of European ancestry and 2747007 SNPs. Data had been age-adjusted and details of the original study is available elsewhere [30].

The study selected for height with the GWAS-ID “ieu-a-97” report summary information from a meta-analysis of 46 studies spanning 73137 females of European ancestry and 2748546 SNPs. Data had been age-adjusted and further information of the study is published elsewhere [30].

For BMI, the study with the GWAS-ID “ieu-a-974” was selected, which report summary data from a large-scale meta-analysis of 82 GWASs over 171977 females of European ancestry and 2494613 SNPs. Data had been age-adjusted and details are given elsewhere [31].

For WC and HC, we selected studies with the GWAS-IDs “ieu-a-63” and “ieu-a-51” respectively, which report pooled statistics from large-scale meta-analyses of 127997 females of European ancestry and 2444355 SNPs. Data had been adjusted for age and other study-specific covariates and details are available elsewhere [32].

The study with the GWAS-ID “ieu-a-75” was selected for WHR, which report pooled results from a meta-analysis encompassing 118003 females of European ancestry and 2466102 SNPs. Data had been adjusted for age and other study-specific covariates and details are available elsewhere [32].

### 2.2. Data source for PCOS

In order to avoid sample-overlapping with exposures, we selected PCOS GWAS summary statistics from a different source i.e. the FinnGen cohort on the IEU Open GWAS Project database. The FinnGen study entails a growing repository of genomic and clinical data emanating from a nationwide network of Finnish biobanks (https://finngen.gitbook.io/documentation/). We used the GWAS with the specific ID “finn-a-E4_POCS” (E4_POCS is the FinnGen phenocode for PCOS) which consisted of 96391 samples (219 cases; 96172 controls) and 16152119 genotyped SNPs in total (**Table 1**). All PCOS cases were clinically diagnosed from hospital discharge registries and cause of death registries using female-specific clinical endpoints (ICD-10: E282, ICD-8: 25690).

### 2.3. Selection of genetic variants as IVs

The SNPs selected as IVs need to fulfil three key assumptions for MR to yield valid results: (1) strongly associated with the exposure (relevance); (2) not associated with the outcome due to confounding (independence); (3) affect the outcome only through the exposure (exclusion restriction). In order to fulfil the first criterion of relevance, we selected biologically and statistically plausible SNPs at a genome-wide significance threshold of *p* < 5e-08. We assessed the statistical power of individual SNPs and the potential weak instrument bias via F-statistic, as defined below.

*F* = *v*^2^ x (*n* - 2)/(1 - *v*^2^), in which *v*^2^ indicates the variance of the exposure phenotype attributable to a given SNP and n stands for the sample size. Variance estimates were calculated using the following formula:

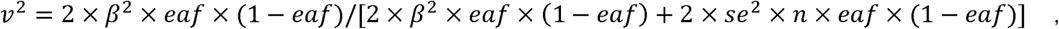

in which *β* denotes the per-allele effect size of the association between a given SNP and the exposure phenotype, *eaf* stands for the effect allele frequency, and *se* is the standard error of *β* [33]. Weak instrument bias may ensue when *F* < 10 [34].

We minimized confounding through the inclusion of female-only cohorts of European ancestry from a single consortium while the summary data had been adjusted for age and other study specific covariates. The assumption of exclusion restriction may be violated in the presence of horizontal pleiotropy, especially unbalanced (directional) horizontal pleiotropy – where one or more IVs exert a net effect on the outcome via a pathway not involving the exposure, biasing the MR estimate. In order to overcome horizontal pleiotropy, we conducted robust MR methods, post-hoc pleiotropy analyses, heterogeneity tests and outlier analyses [35], the details of which are provided later in the manuscript.

As the absence of linkage disequilibrium (LD) is a prerequisite for most 2SMR methods, we performed clumping to prune statistically plausible IVs (*p* < 5e-08), selecting those SNPs with LD-R^2^ < 0.001 and clumping-distance > 10000 kb. When any exposure SNP was not present in the outcome data, we included proxy SNPs via LD tagging, with following specifications: minimum LD-R^2^ value = 0.8; minor allele frequency threshold for aligning palindromic SNPs = 0.3. We performed allele harmonization by aligning strands for all SNPs including palindromes, to ensure that the effects of the SNPs on the exposure correspond to the same allele as their effects on the outcome.

### 2.4. 2SMR analyses

We applied four MR methods i.e. inverse variance weighted (IVW) method using multiplicative random effects model as the primary analysis along with three robust methods: MR-Egger (MRE), weighted median (WME), and weighted mode (WMO) [35].

The IVW method is the recommended main MR method to be used with summarized data and multiple, uncorrelated genetic variants, since it is the most efficient approach in the presence of valid IVs, yielding causal estimates that are accounted for heterogeneity [35]. This method pools Wald ratio estimates for individual SNPs using inverse of the variance as weights.

The MRE method provides an asymptotically consistent causal effect measure adjusted for horizontal pleiotropy by pooling individual SNP-specific Wald ratios via an adapted Egger regression. In fact, its regression intercept is an estimate of the net pleiotropic effect which can therefore be used to assess horizontal pleiotropy. It allows for invalid IVs, provided the Instrument Strength Independent of Direct Effect (INSIDE) assumption (i.e. instrument strengths are independent of horizontal pleiotropic effects) holds true. However, this method is underpowered in the presence of relatively homogeneous SNP-exposure effect sizes, susceptible to regression dilution bias, and the causal estimation is heavily affected by outliers [35-37].

Using the weighted median of Wald ratios, the WME method produces an asymptotically consistent estimate of the causal effect, provided 50% or more of the variants are valid IVs that do not violate the exclusion restriction criterion [35]. The WMO method clusters SNPs into groups based on the similarity of their individual ratio estimates, calculates the inverse variance weighted number of SNPs in each cluster, and produces a causal estimate based on the cluster having the largest weighted number of SNPs [38]. Both WME and WMO methods require some genetic variants to be valid instruments and are robust to outliers [35].

Results from 2SMR analyses were summarized in tabular format, along with odds ratios (OR) and 95% confidence intervals (CI) for all β estimates.

### 2.5. Method comparison plots

Scatter plots and trend lines pertaining to different MR methods were generated for each anthropometric exposure-outcome analysis. Slopes and directions of trend lines represent the magnitudes and directions of causal estimates, respectively.

### 2.6. Single SNP analyses

Causal effects of each SNP were determined individually and were visualized in forest plots along with pooled estimates using all SNPs, under IVW and MRE methods. With respect to each exposure, we investigated for the presence of any significantly causally associated individual SNPs at Bonferroni multiple testing corrected *p*-value thresholds.

### 2.7. Leave-one-out analyses

We conducted leave-one-out sensitivity analyses to assess whether causal estimates were significantly influenced by a single SNP. Wald ratios from IVW-MR analyses conducted excluding each SNP as well as pooled IVW-MR estimates encompassing all SNPs were visualized in forest plots.

### 2.8. Heterogeneity analyses

Heterogeneity is a measure of the consistency of the causal estimate across all SNPs whereby lower heterogeneity is indicative of a reliable MR estimate. We assessed heterogeneity via Cochran’s Q statistic and associated *p*-values. Funnel plots were also generated to visually assess heterogeneity. Lower values on the y-axis denote less precise estimates while those with increasing precision tend to ‘funnel’ in. Larger distributions suggest heterogeneity which may have resulted from horizontal pleiotropy whereas asymmetric distributions are indicative of unbalanced horizontal pleiotropy.

### 2.9. Analysis of horizontal pleiotropy and outliers

The MRE regression intercept is an estimator of the magnitude of horizontal pleiotropy [36]. For each anthropometric exposure, we calculated the MRE intercept, its standard error, and directionality *p*-value. The Mendelian Randomization Pleiotropy RESidual Sum and Outlier (MRPRESSO) method presents a unified framework to assess pleiotropy and outliers via a three-step process: identification of pleiotropy and outliers (MRPRESSO global test); rectification of pleiotropy by removing outliers (MRPRESSO outlier test); analysis of the distortion in the causal estimate before and after removing outliers (MR-PRESSO distortion test) [39]. We applied the MRPRESSO method to evaluate pleiotropy and outliers with respect to all exposure-outcome associations. Radial plots have been proposed as an improved strategy for visualizing outliers in 2SMR analyses [40]. We generated radial plots for all exposure-outcome associations that were investigated with 2SMR in this study.

All analyses were conducted in R (version 4.1.2) [41], using “TwoSampleMR” [42], “MRPRESSO” [39], and “RadialMR” [40] packages.

### 2.10. Exploring genetic architectures underlying causality

For significant causal associations, we predicted nearest gene(s)/transcriptional start site(s) ascribed to corresponding SNPs in harmonized datasets, using the Open Targets Genetics Portal [43]. In order to identify shared and non-shared genetic architectures underlying these significant causal associations, we explored overlapping and non-overlapping SNPs and genes across exposures.

## 3. RESULTS

### 3.1. Genetic variants selected as IVs

Harmonized datasets containing details of the SNPs selected as IVs for each exposure are presented in **S2 Table**. Nine SNPs underlying the weight-PCOS association were identified, which comprised 3 proxies and no palindromes. We included 43 SNPs with respect to the height-PCOS association, which consisted of 23 proxies and 2 palindromes. Thirty-one SNPs were identified with respect to the BMI-PCOS association, which comprised 16 proxies and 2 palindromes. With respect to WC-PCOS and HC-PCOS associations, we identified 15 identical SNPs, which comprised 7 proxies and no palindromes. We identified 18 SNPs underlying the WHR-PCOS association, which consisted of 7 proxies and no palindromes. The F-statistic of all IVs across the exposures was > 29, indicating no substantial weak instrument bias.

### 3.2. 2SMR results

Results from 2SMR analyses are presented in **Table 2**. Three exposures, namely, BMI, WC, and HC, were significantly causally associated with PCOS.

**Table 2:** Results from two-sample Mendelian randomization analyses on the causality of anthropometric markers associated with polycystic ovarian syndrome.

| Anthropometric marker | Method | No: of SNPs | $\beta$ (SE) | 95% CI of $\beta$ | *p*-value | OR | 95% CI of OR |
| --- | --- | --- | --- | --- | --- | --- | --- |
| Weight | MR Egger | 9 | 4.53 (2.83) | -1.02, 10.07 | 0.1537 | 92.37 | 0.36, 23649.56 |
|  | Weighted median | 9 | 1.45 (0.95) | -0.43, 3.33 | 0.1256 | 4.26 | 0.65, 28.01 |
|  | IVW | 9 | 1.08 (0.71) | -0.31, 2.47 | 0.1292 | 2.94 | 0.73, 11.85 |
|  | Weighted mode | 9 | 1.85 (1.25) | -0.79, 4.50 | 0.1767 | 6.37 | 0.45, 89.68 |
| Height | MR Egger | 43 | -2.68 (1.79) | -6.19, 0.82 | 0.1416 | 0.07 | 0.002, 2.28 |
|  | Weighted median | 43 | -0.33 (0.56) | -1.43, 0.77 | 0.5619 | 0.72 | 0.24, 2.17 |
|  | IVW | 43 | 0.06 (0.43) | -0.79, 0.92 | 0.8849 | 1.06 | 0.45, 2.50 |
|  | Weighted mode | 43 | -1.34 (1.20) | -3.57, 0.89 | 0.2703 | 0.26 | 0.03, 2.44 |
| **BMI** | MR Egger | 31 | 2.50 (1.39) | -0.23, 5.23 | 0.0835 | 12.20 | 0.79, 187.88 |
|  | **Weighted median** | **31** | **1.94 (0.75)** | **0.44, 3.43** | **0.0096** | **6.94** | **1.55, 31.01** |
|  | IVW | 31 | 1.71 (0.53) | 0.68, 2.75 | 0.0012 | 5.55 | 1.97, 15.65 |
|  | **Weighted mode** | **31** | **1.98 (0.90)** | **0.04, 3.91** | **0.0365** | **7.24** | **1.04, 50.19** |
| **WC** | MR Egger | 15 | 2.15 (3.03) | -3.80, 8.10 | 0.4910 | 8.59 | 0.02, 3284.49 |
|  | **Weighted median** | **15** | **2.70 (1.13)** | **0.59, 4.81** | **0.0166** | **14.88** | **1.80, 122.91** |
|  | IVW | 15 | 1.92 (0.88) | 0.18, 3.65 | 0.0302 | 6.79 | 1.20, 38.37 |
|  | **Weighted mode** | **15** | **3.20 (1.66)** | **0.06, 6.34** | **0.0498** | **24.56** | **1.06, 569.79** |
| **HC** | MR Egger | 15 | 2.15 (3.03) | -3.80, 8.09 | 0.4912 | 8.57 | 0.02, 3271.61 |
|  | **Weighted median** | **15** | **2.70 (1.12)** | **0.59, 4.81** | **0.0159** | **14.88** | **1.80, 122.97** |
|  | IVW | 15 | 1.91 (0.88) | 0.18, 3.64 | 0.0303 | 6.78 | 1.20, 38.28 |
|  | **Weighted mode** | **15** | **3.20 (1.65)** | **0.17, 6.24** | **0.0495** | **24.56** | **1.18, 511.07** |
| **WHR** | MR Egger | 18 | 1.16 (3.71) | -6.12, 8.43 | 0.7592 | 3.18 | 0.002, 4607.52 |
|  | Weighted median | 18 | -0.26 (0.93) | -2.09, 1.57 | 0.7779 | 0.77 | 0.12, 4.80 |
|  | IVW | 18 | -0.64 (0.69) | -2.00, 0.71 | 0.3492 | 0.52 | 0.14, 2.02 |
|  | Weighted mode | 18 | -0.11 (1.27) | -2.84, 2.61 | 0.9314 | 0.89 | 0.06, 13.66 |

**BMI = body mass index; HC = hip circumference; IVW = inverse variance weighted method; WC = waist circumference; WHR = waist to hip ratio**

According to the IVW method, a unit (1 kg/m^2^) increase in BMI was associated with a significant increase in PCOS (OR = 5.55; 95% CI = 1.97, 15.65; *p* = 0.0012). As per the WME model, a unit increase in BMI yielded an OR of 6.94 (95% CI = 1.55, 31.01; *p* = 0.0096). As revealed by the WMO method, a unit increase in BMI conveyed an OR of 7.24 (95% CI = 1.04, 50.19; *p* = 0.0365).

The IVW method revealed that WC was significantly associated with a higher risk of PCOS (OR = 6.79; 95% CI = 1.20, 38.37; *p* = 0.0302). As per the WME method, a unit (1cm) increase in WC significantly increased PCOS risk (OR = 14.88; 95% CI = 1.80, 122.91; *p* = 0.0166). According to the WMO method, a unit increase in WC conveyed an OR of 24.56 (95% CI = 1.06, 569.79; *p* = 0.0498).

As revealed by the IVW method, a unit (1cm) increase in HC was significantly associated with a higher risk of PCOS (OR = 6.78; 95% CI = 1.20, 38.28; *p* = 0.0303). According to the WME method, a unit increase in HC was associated with a significant increase in PCOS (OR = 14.88; 95% CI = 1.80, 122.97; *p* = 0.0159). As per the WMO method, a unit increase in HC conveyed an OR of 24.56 (95% CI = 1.18, 511.07; *p* = 0.0495).

However, MRE regression estimates did not achieve statistical significance with BMI, WC, and HC as exposures. Also, weight, height, and WHR were not significantly causally associated with PCOS in any 2SMR analyses.

### 3.3. Method comparison plots

As shown in **Figure 1**, scatter plots and trend lines pertaining to the three significant exposures (BMI, WC, HC) demonstrate positive causal associations with PCOS, according to different MR methods. Scatter plots and regression lines for the three non-significant exposures (weight, height, WHR) are presented in **S1 Figure**.

**Figure 1:**
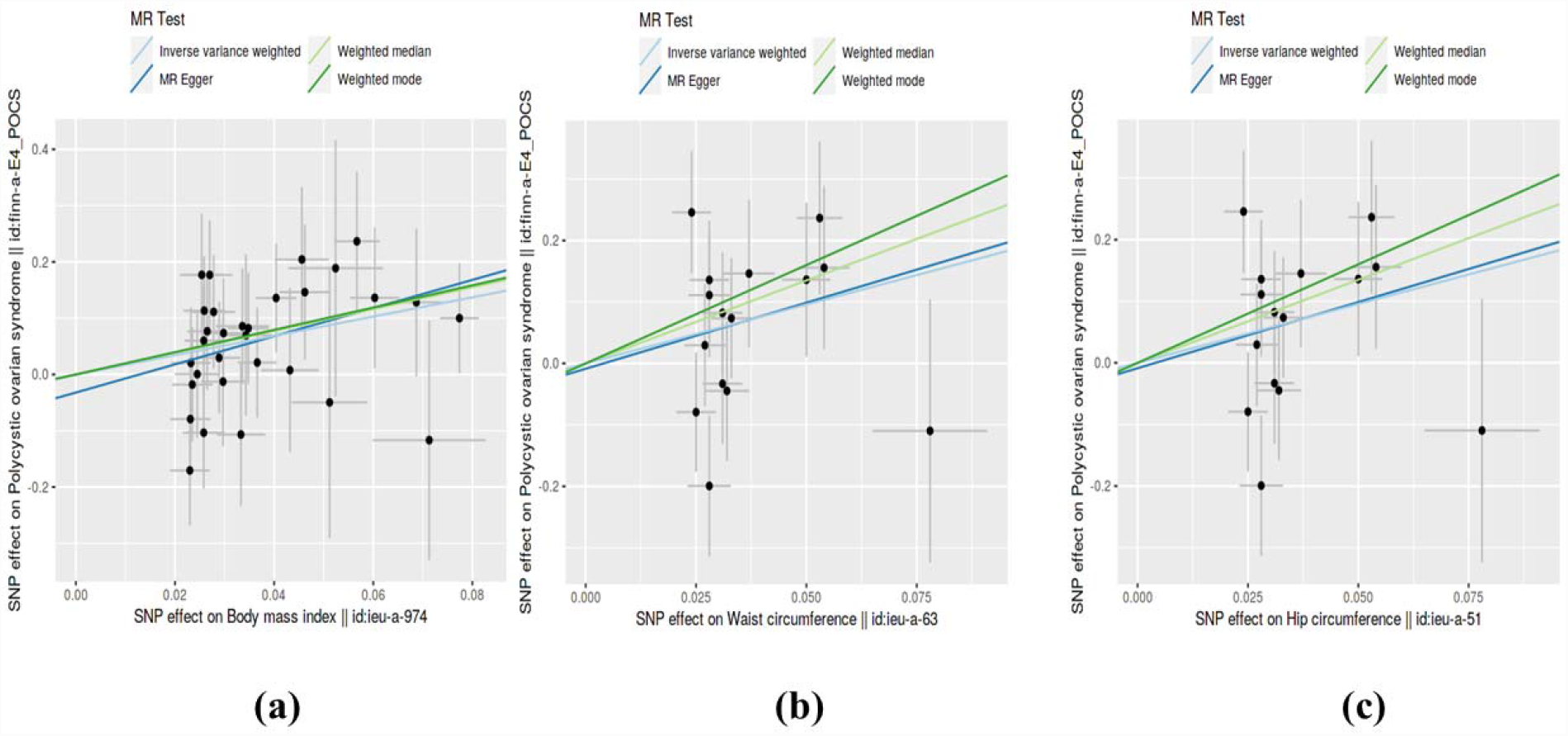
Scatter plots illustrating the distribution of individual ratio estimates of the three significant exposures: (a) body mass index (b) waist circumference (c) hip circumference, with polycystic ovarian syndrome as the outcome. Trend lines from the four different Mendelian randomization methods employed indicating the positive causal associations, are also included in each scatter plot.

### 3.4. Single SNP analyses

Results from single SNP analyses are provided in **S3 Table**. Forest plots depicting single SNP analyses and pooled causal estimates as per IVW and MRE methods for significant exposures are presented in **Figure 2** while corresponding forest plots for non-significant exposures are given in **S2 Figure**. According to multiple testing corrected *p*-value thresholds, none of the SNPs were individually associated with PCOS in all exposure data: weight (*p* > 5.56 × 10^−3^, 0.05/9); height (*p* > 1.16 × 10^−3^, 0.05/43); BMI (*p* > 1.61 × 10^−3^; 0.05/31); WC and HC (*p* > 3.33 × 10^−3^; 0.05/15); WHR (*p* > 2.78 × 10^−3^; 0.05/18).

**Figure 2:**
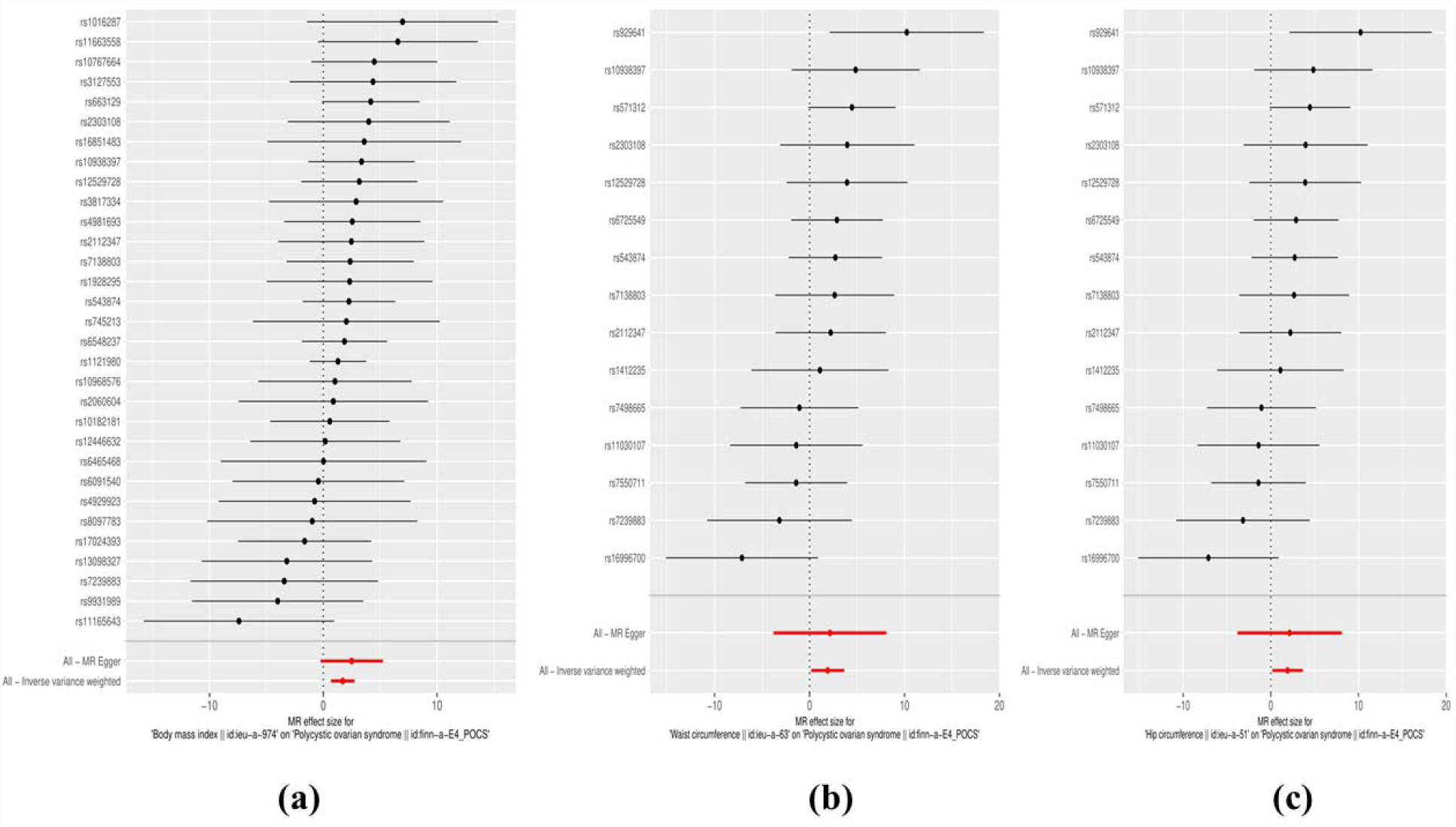
Forest plots of the three significant exposures: (a) body mass index (b) waist circumference (c) hip circumference, against polycystic ovarian syndrome as the outcome. Effects of individual SNPs and pooled estimates from MR-Egger- and inverse variance weighted methods are visualized.

### 3.5. Leave-one-out analyses

Results from leave-one-out analyses are presented in **S4 Table**. Leave-one-out analysis plots and pooled causal estimates as per IVW-MR method (for comparison) for significant exposures are presented in **Figure 3** while corresponding leave-one-out analysis plots for non-significant exposures are provided in **S3 Figure**. According to Bonferroni multiple testing corrected *p*-value thresholds, none of the leave-one-out sensitivity analyses yielded statistical significance, indicating robustness of causal estimates: weight (*p* > 5.56 × 10^−3^, 0.05/9); height (*p* > 1.16 × 10^−3^, 0.05/43); BMI (*p* > 1.61 × 10^−3^; 0.05/31); WC and HC (*p* > 3.33 × 10^−3^; 0.05/15); WHR (*p* > 2.78 × 10^−3^; 0.05/18).

**Figure 3:**
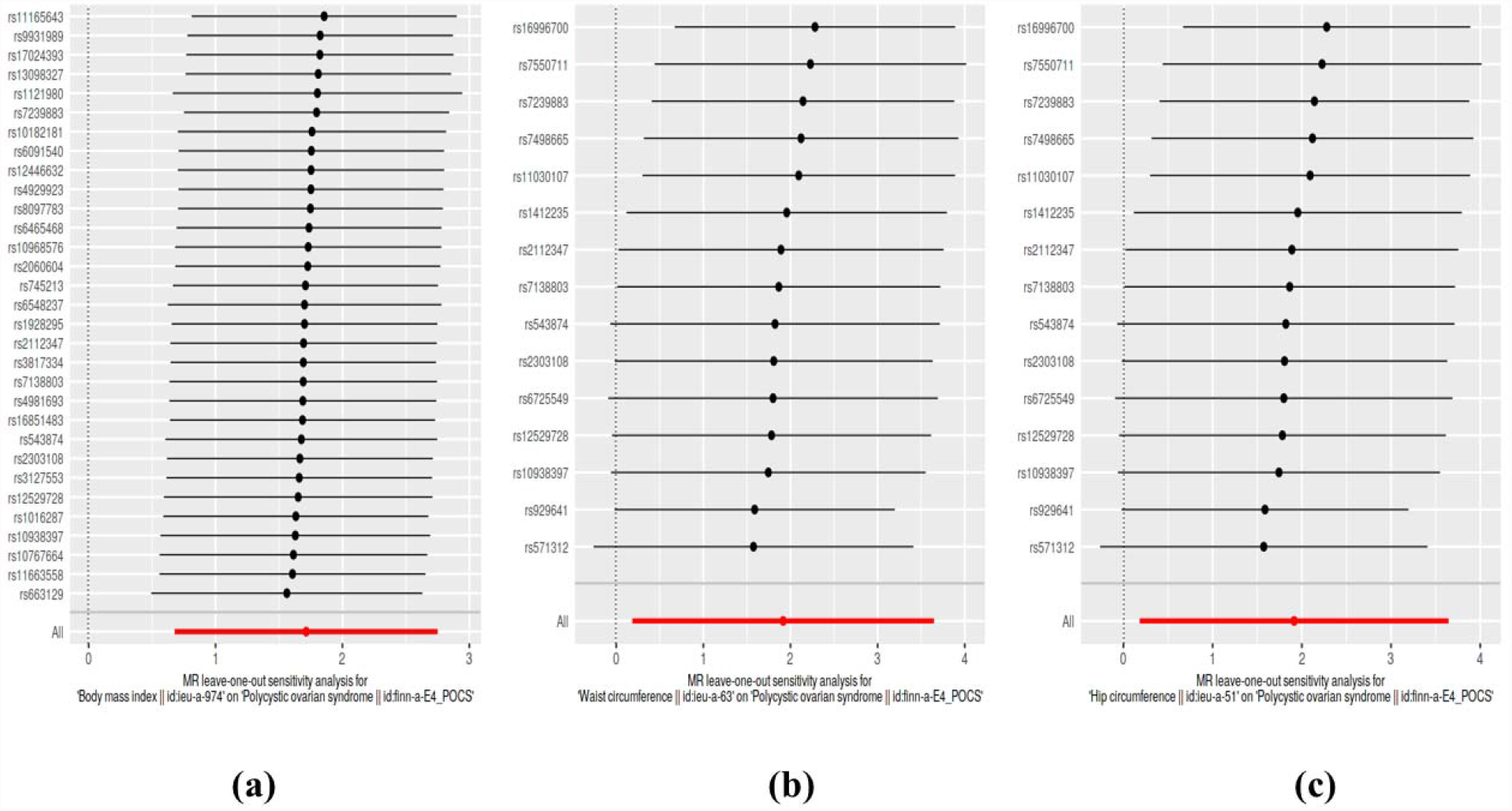
Leave-one-out sensitivity analysis plots of the three significant exposures: (a) body mass index (b) waist circumference (c) hip circumference, against polycystic ovarian syndrome as the outcome. A given dark point indicates the effect measure from inverse variance weighted Mendelian randomization analysis excluding that specific SNP. The red lines indicate pooled analyses encompassing all SNPs via IVW-MR method (drawn for comparison).

### 3.6. Heterogeneity analyses

Results from heterogeneity analyses (Cochran’s Q- and *p*-values) are summarized in **Table 3**. We did not observe statistically significant heterogeneity in any 2SMR analyses (*p* > 0.05). Funnel plots for the significant exposures are illustrated in **Figure 4** while those for non-significant exposures are provided in **S4 Figure**. In conformity with non-significant findings from heterogeneity analyses, no asymmetric distributions indicating directional horizontal pleiotropy or large spreads suggesting considerable heterogeneity could not be discerned in funnel plots.

**Table 3:** Heterogeneity statistics of two-sample Mendelian randomization analyses.

| **Anthropometric marker** | **Method** | **Q value** | **Degrees of freedom** | **p-value** |
| --- | --- | --- | --- | --- |
| Weight | MR Egger | 6.589 | 7 | 0.4729 |
|  | IVW | 8.172 | 8 | 0.4169 |
| Height | MR Egger | 56.54 | 41 | 0.0538 |
|  | IVW | 56.98 | 42 | 0.0544 |
| BMI | MR Egger | 20.30 | 29 | 0.8835 |
|  | IVW | 20.67 | 30 | 0.8980 |
| Waist circumference | MR Egger | 16.86 | 13 | 0.2060 |
|  | IVW | 16.86 | 14 | 0.2635 |
| Hip circumference | MR Egger | 16.84 | 13 | 0.2065 |
|  | IVW | 16.85 | 14 | 0.2641 |
| Waist to hip ratio | MR Egger | 12.79 | 16 | 0.6881 |
|  | IVW | 13.03 | 17 | 0.7339 |

**BMI = body mass index; IVW = inverse variance weighted method**

**Figure 4:**
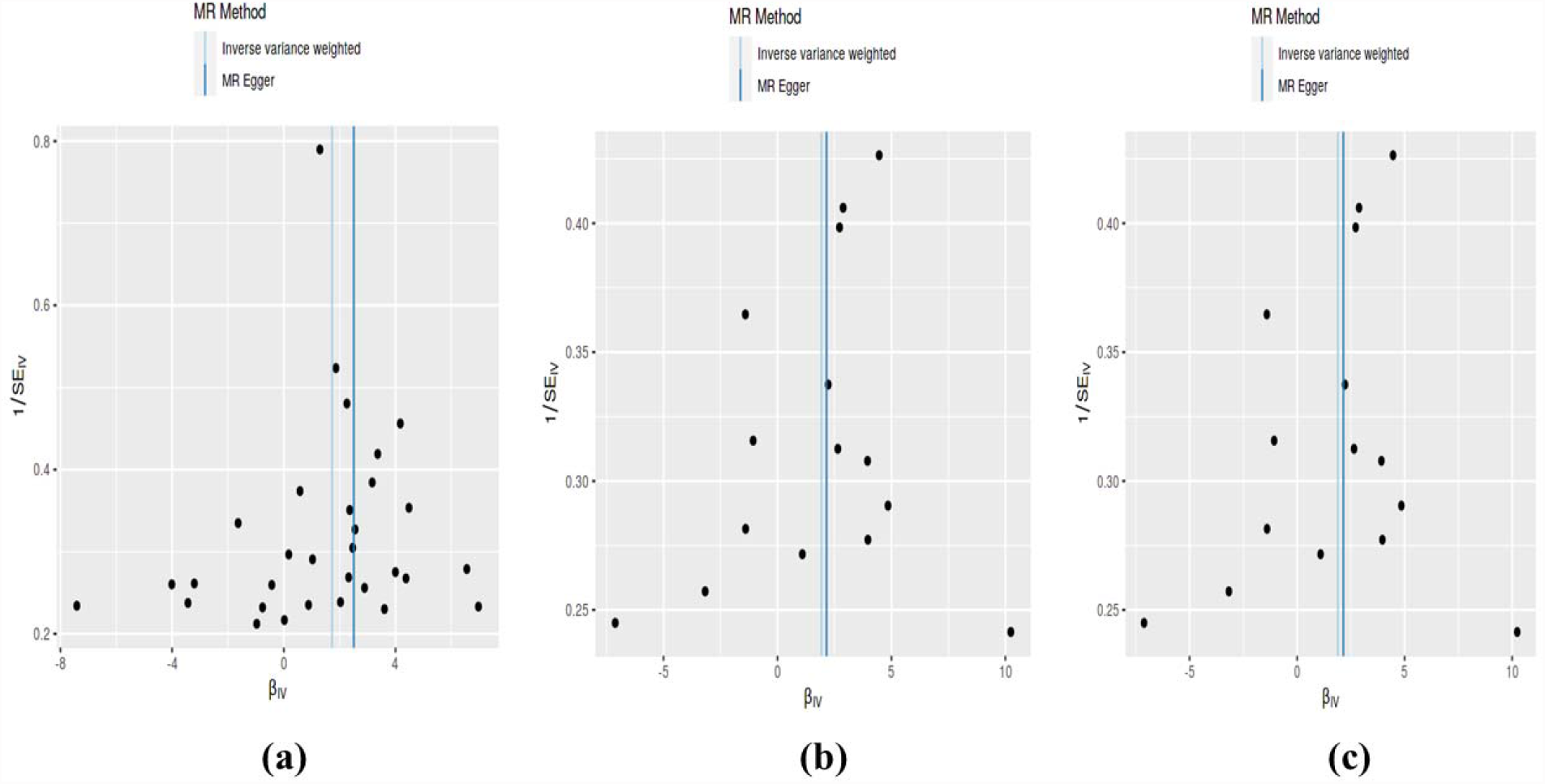
Funnel plots of the three significant exposures: (a) body mass index (b) waist circumference (c) hip circumference, against polycystic ovarian syndrome as the outcome.

### 3.7. Horizontal pleiotropy and outliers

We present the results of horizontal pleiotropy analyses via MRE regression intercepts and directionality *p*-values in **Table 4**. These analyses revealed that there was no significant horizontal pleiotropy (*p* > 0.05). As shown in **S5 Table**, congruent findings were obtained from MRPRESSO analyses which confirmed the absence of outliers and no significant horizontal pleiotropy (global test *p*-values > 0.05). Radial plots for significant exposures are provided in **Figure 5** while those for non-significant exposures are given in **S5 Figure**. All radial plots illustrate the absence of outliers.

**Table 4:** Horizontal pleiotropy statistics of two-sample Mendelian randomization analyses.

| **Anthropometric marker** | **Egger regression intercept** | **Standard error** | **Directionality p-value** |
| --- | --- | --- | --- |
| Weight | -0.18 | 0.15 | 0.249 |
| Height | 0.13 | 0.081 | 0.122 |
| Body mass index (BMI) | -0.032 | 0.053 | 0.547 |
| Waist circumference (WC) | -0.0086 | 0.11 | 0.937 |
| Hip circumference (HC) | -0.0086 | 0.11 | 0.937 |
| Waist to hip ratio (WHR) | -0.065 | 0.13 | 0.628 |

**Figure 5:**
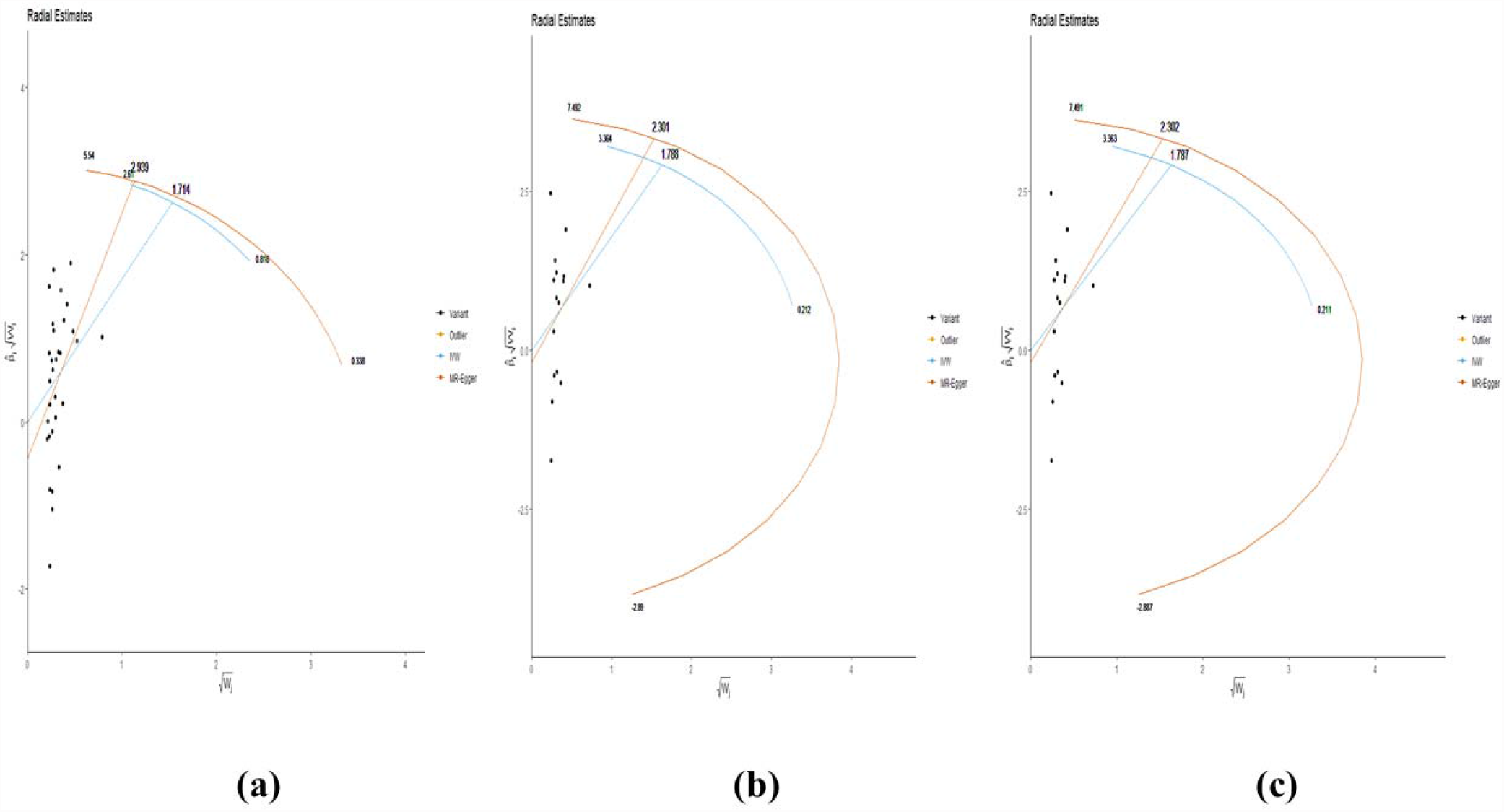
Radial plots of the three significant exposures (a) body mass index (b) waist circumference (c) hip circumference. No significant outliers were detected.

### 3.8. Genetic architectures underlying causality

**Table 5** presents SNPs and nearest genes/TSSs underlying the three significant causal associations: BMI, WC and HC versus PCOS. Notably, both WC and HC had the same 15 SNPs underlying their significant causal associations with PCOS. There were 31 SNPs underlying the association between BMI and PCOS. Seven common SNPs (*rs10938397, rs12529728, rs2112347, rs2303108, rs543874, rs7138803, rs7239883*) were observed across these associations. We identified 34 genes underlying the association between BMI and PCOS and 19 genes underlying the associations between WC/HC and PCOS. Of these, 17 genes (*ATP2A1, BCDIN3D, BDNF, FANCL, GNPDA2, LINGO2, MC4R, NCKAP5L, POC5, RIT2, SAE1, SEC16B, SH2B1, TFAP2B, TMEM18, ZC3H4, ZFP64*) were shared across the three significant associations. Seventeen genes (*ADCY3, AMPD2, BEND5, CADM2, FOXG1, FTO, GPRC5B, HNF4G, MTCH2, NPC1, PDK4, PTBP2, RASA2, RPGRIP1L, SKOR1, STK33, TLR4*) were exclusive to the association between BMI and PCOS while two genes (*GPR61, VRK2*) were exclusive to the association between WC/HC and PCOS.

## 4. DISCUSSION

To our knowledge, this is one of the first 2SMR studies to comprehensively and systematically analyze causal associations between anthropometric markers of both general- and central-obesity in large-sample, female-only cohorts of European ancestry and report shared and non-shared genetic architectures with respect to their causality in ensuing PCOS. We found that both central- and general-obesity potentially cause PCOS, as signified by the three significant anthropometric markers – BMI, WC, and HC. With respect to general obesity, our findings are congruent with previous MR analyses which reported positive causal associations of BMI with PCOS [25-28]. With regard to central obesity, we observed strong positive causal associations of WC and HC with PCOS, as indicated by relatively large OR values (6.78-24.56).

**Table 5:**
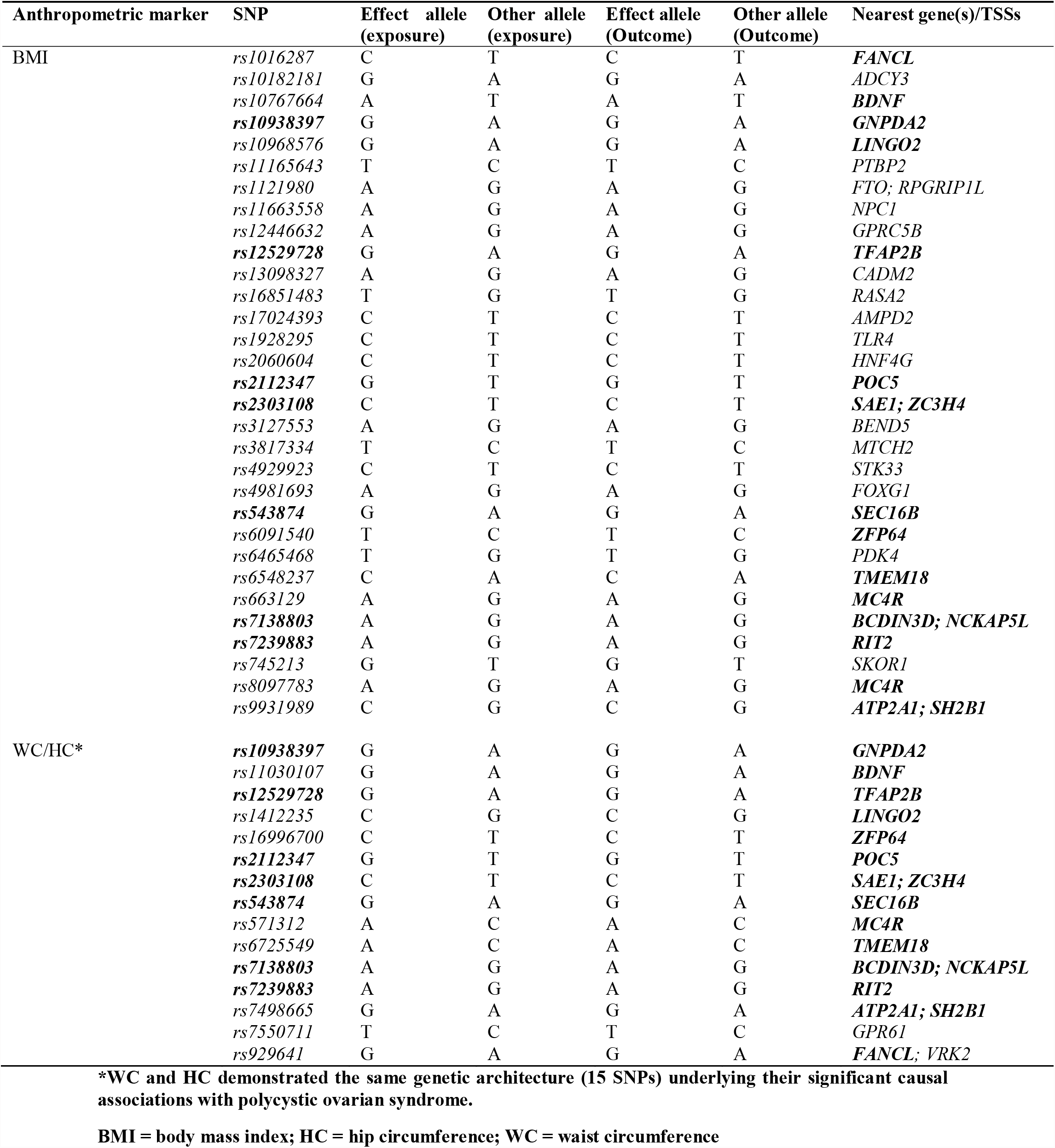
Genetic variants (SNPs) and nearest genes/transcriptional start sites (TSSs) underlying the significant causal associations.

We observed consistent and significant causal associations from IVW method and two robust methods (WME and WMO methods) with regard to BMI, WC, and HC which was reassuring. MRE method did not yield significant results, nonetheless, which could perhaps be due to its lack of precision in the presence of relatively homogeneous variant-exposure effect sizes [35], given that we applied stringent eligibility criteria to select only highly uncorrelated and independent IVs (LD-R^2^ = 0.001). We ensured minimal overlap between exposure-outcome samples – a key requirement for reducing bias in 2SMR estimates [14]. In fact, non-substantial sample overlaps, especially involving controls, are even permissible, and would not evoke bias in 2SMR studies [44]. No confounding by ancestry entailed as all participants were of European ancestry. However, this may have led to a lack of generalizability of our findings and transethnic validations via MR in independent cohorts would be required. Further, there was no weak instrument bias as all IVs had F > 10. Robustness of our findings was underscored by a range of sensitivity analyses, which uncovered no substantial heterogeneity, horizontal pleiotropy, and outliers. While the use of summarized public data has advantages such as transparency, reproducibility, and the greater ability to detect causal associations due to large samples, it has a few shortcomings as well. For example, we could not conduct subgroup analyses, rule out collider bias due to over-adjusted summarized association estimates, or assess potentially non-linear associations of obesity with PCOS, for which individual-level data are required [35]. The relatively small number of PCOS cases in the outcome GWAS (N=219) might be a limiting factor, given the community prevalence of PCOS > 8% [2], as opposed to the substantially lower prevalence of clinically-diagnosed cases (∼0.2%) in the Finnish cohort used in the present study. However, these are women with overt PCOS presenting to the clinicians with common, cardinal clinical features including obesity and predominant PCOS phenotypes, as confirmed by ICD codes. As we uncovered potential causalities despite the small number of cases, stronger findings with larger causal estimates might possibly be obtained from independent cohorts containing a larger number of women with PCOS, which need to be investigated in future studies. Although we minimized confounding with female-only cohorts of European ancestry and summary estimates adjusted for multiple variables, there may still have been residual confounding influencing our causal estimates toward or away from the null.

Mechanisms underlying the causal effects of central-and general obesity on PCOS are not fully understood and need to be investigated. Towards this end, the SNPs and neighboring genes underlying causal associations uncovered in the present study may be insightful. We observed a considerable degree of overlap (7 SNPs; 17 genes) across significant causal findings, perhaps indicating similar genetically-driven causal mechanisms. However, there were SNPs and genes that were exclusive to each causal association as well, suggesting unique, exposure-specific pathways may also participate in PCOS pathogenesis. This suggests that differences may exist in the genetic basis of PCOS among women with central obesity alone, as compared to women with both general- and central obesity. Of the identified genes in our study, *FTO, GNPDA2, MC4R, MTCH2, SH2B1, TMEM18* are unequivocally regarded as obesity genes associated with PCOS, as supported by contemporary evidence [45]. A gene set rather similar to those found in the present study (*ADCY3, BCDIN3D, BDNF, CADM2, FTO, GNPDA2, GPRC5B, HNF4G, LINGO2, MC4R, MTCH2, POC5, PTBP2, RASA2, SEC16B, TFAP2B, TLR4, TMEM18, ZC3H4, ZFP64*) was underlying the causal association between BMI and PCOS in a previous MR study as well [25].

There is a formidable body of evidence supporting *FTO* gene’s association with both obesity and PCOS. While *FTO* is, in fact, the first locus identified as unequivocally associated with adiposity [46], a meta-analysis revealed a direct association between *FTO* variant and PCOS risk, independent of BMI [47]. Both *FTO* and *MC4R* gene variants are associated with obesity in PCOS [48] while observational evidence indicates a direct role of the interaction between *FTO* and *MC4R* polymorphisms in the development of PCOS [49]. *TLR4* and toll-like receptor genes in general contribute to the development of chronic low-grade inflammation as well as insulin resistance and hyperandrogenism observed in PCOS [50] whereas a MR study revealed essential causal roles of systemic inflammatory regulators, especially cytokines, in the pathogenesis of PCOS [51]. However, mechanisms by which many other genes found to be underlying the causal associations in the present study contribute to the development of PCOS, are not completely known.

Current evidence indicates that obesity contributes to the onset and progression of PCOS through multiple mechanisms including the escalation of insulin resistance and compensatory hyperinsulinemia which in turn leads to augmented adipogenesis and reduced lipolysis, sensitization of thecal cells to luteinizing hormone and intensification of functional ovarian hyperandrogenism by increasing ovarian androgen synthesis, and elevation of inflammatory adipokines which in turn leads to upregulation of insulin resistance and adipogenesis [52]. Indeed, obesity contributes to the pathogenesis of PCOS via multiple mechanisms that encompass the three cardinal clinical manifestations of PCOS, namely, hyperandrogenism, reproductive-, and metabolic dysfunction [53]. Moreover, our findings allude to the concept of secondary PCOS, which has only emerged in recent years. It has been proposed that adiposity and hyperinsulinemia (exogenous in type 1- and endogenous in type 2-diabetes) cause PCOS [54]. Recent research on polygenic risks scores has also demonstrated that both men and women manifest similar metabolic features of PCOS and that the presence of ovaries is neither essential nor required for PCOS while obesity features prominently [55]. Genetic research is providing greater insights into the mechanisms and nature of this complex polygenic disorder.

Lastly, lean PCOS phenotypes which are less prevalent than obese or overweight PCOS phenotypes, may have different underlying genetic architectures and pathogenic mechanisms and may need to be investigated separately. Given the differences in hormonal and metabolic profiles, treatment strategies and outcomes between obese- and non-obese PCOS phenotypes, it has been proposed to incorporate obesity as a clinical sign for classifying PCOS phenotypes [56].

Our findings indicating the causality of obesity and adiposity in PCOS have clinical and public health implications. These findings highlight the need to address obesity prevention at a population level through successful fiscal policies and statutory regulations [57]. Our findings are also congruent with recommendations from current international guidelines on PCOS, which underscore the prevention of excess weight gain in all women with PCOS, alongside lifestyle interventions focused on diet, weight loss, and physical activity for women with PCOS with overweight and obesity. As in the standard clinical management of obesity, pharmacotherapy and other treatments may also be appropriate [52].

## 5. CONCLUSIONS

In this study, we revealed that both central- and general obesity potentially cause PCOS, via 2SMR analyses using female-only, large-sample cohorts of European ancestry. Findings provide insight into the mechanisms underpinning this common and complex condition. They also support international guidelines and underscore the importance of addressing obesity and adiposity for the prevention and management of PCOS.

## Data Availability

All data produced are available online at: https://gwas.mrcieu.ac.uk/

## DECLARATIONS

### Ethics approval

Not required being an analysis of publicly available, deidentified, summarized data.

### Author contributions

Kushan De Silva: Conceptualization, Data Curation, Formal Analysis, Funding Acquisition, Investigation, Methodology, Project Administration, Resources, Software, Visualization, Writing – Original Draft Preparation, Writing – Review & Editing

Ryan T. Demmer: Conceptualization, Methodology, Project administration, Resources, Supervision, Validation, Writing – review & editing

Daniel Jönsson: Conceptualization, Methodology, Supervision, Validation, Writing – review & editing

Aya Mousa: Conceptualization, Methodology, Supervision, Validation, Writing – review & editing

Helena Teede: Methodology, Supervision, Validation, Writing – review & editing

Andrew Forbes: Methodology, Supervision, Validation, Writing – review & editing

Joanne Enticott: Conceptualization, Methodology, Project administration, Resources, Supervision, Validation, Writing – review & editing

### Availability of data and material

All data used in this study are freely available in the IEU GWAS database at: https://gwas.mrcieu.ac.uk/

## SUPPORTING INFORMATION CAPTIONS

**S1 Table. STROBE MR checklist**

**S2 Table. Harmonized data**

**S3 Table. Single SNP analysis**

**S4 Table. Leave-one-out analysis**

**S5 Table. MRPRESSO analysis**

**S1 Fig. Scatter plots of ratio estimates for the three non-significant exposures**

**S2 Fig. Forest plots of the three non-significant exposures**

**S3 Fig. Leave-one-out sensitivity analysis plots of the three non-significant exposures**

**S4 Fig. Funnel plots of the three non-significant exposures**

**S5 Fig. Radial plots of the three non-significant exposures**

