## Supplementary material for "Both general- and central- obesity are causally associated with polycystic ovarian syndrome: Findings of a Mendelian randomization study": S1 Table

### Strobe-MR checklist

| **Item** | **Complete/location** |
| --- | --- |
| 1. **Title and Abstract:** "Mendelian randomization" is named both in the title and the abstract | **Complete** |
| **Introduction** |  |
| 2. **Background:** Explain the scientific background and rationale for the reported study. Is causality between exposure and outcome plausible? Justify why MR is a helpful method to address the study question. | In the third and fourth paragraphs of Introduction, we discuss why central- and general- obesity are plausible causal exposures for PCOS. In the second paragraph, we explain why Mendelian randomization is a reasonable approach to investigate potential causality between central- and general- obesity and PCOS. |
| 3. **Objectives:** State specific objectives clearly, including pre-specified causal hypotheses (if any). | In the 4^th^ and 5^th^ paragraphs of Introduction, we state the objectives of our study and describe pre-specified causal hypotheses. |
| **Methods** |  |
| 4. **Study design and data sources:** Present key elements of study design early in the paper. Consider including a table listing sources of data for all phases of the study. For each data source contributing to the analysis, describe the following:   a) Describe the study design and the underlying population from which it was drawn. Describe also the setting, locations, and relevant dates, including periods of recruitment, exposure, follow-up, and data collection, if available.  b) Give the eligibility criteria, and the sources and methods of selection of participants.  c) Explain how the analyzed sample size was arrived at.  d) Describe measurement, quality and selection of genetic variants.  e) For each exposure, outcome and other relevant variables, describe methods of assessment and, in the case of diseases, the diagnostic criteria used.  f) Provide details of ethics committee approval and participant informed consent, if relevant. | Information about the GWAS studies summarized in Table 1, and the section “GWAS details”.   a) Further information on these aspects is given in each of the original GWAS publications and we have provided references. b) In 2.1. and 2.2. sections of Methods, we have described eligibility criteria, data sources, and methods of selection of cohorts. c) As mentioned in 2.1. and 2.2. sections of Methods, we prioritized GWASs with large sample sizes. While formal sample size calculations were not performed, we assumed that the selected large sample cohorts were adequately powered. d) These aspects are described in 2.3. section of Methods. e) Further information on these aspects is given in each of the original GWAS publications and we have provided references. f) All original GWAS studies had obtained ethics approval and participant informed consent, as stated in respective publications. Since we used publicly available, summarized, and deidentified data from these GWASs, ethics approval was not required for our analysis. |
| 5. **Assumptions:** Explicitly state assumptions for the main analysis (e.g. relevance, exclusion, independence, homogeneity) as well assumptions for any additional or sensitivity analysis. | Described in 2.3. and 2.4. sections of Methods. |
| 6. **Statistical methods main analysis**   Describe statistical methods and statistics used.  a) Describe how quantitative variables were handled in the analyses (i.e., scale, units, model).  b) Describe the process for identifying genetic variants and weights to be included in the  analyses (i.e, independence and model). Consider a flow diagram.  c) Describe the MR estimator, e.g. two-stage least squares, Wald ratio, and related statistics.  Detail the included covariates and, in case of two-sample MR, whether the same covariate set was used for adjustment in the two samples.  d) Explain how missing data were addressed.  e) If applicable, say how multiple testing was dealt with. | All statistical methods and statistics used are described in Methods.  (a) Described in 2.3. and 2.4. sections of Methods.  b) Described in 2.3 and 2.4. sections of Methods.  c) Described in 2.1. and 2.4. sections of Methods.  d) Not applicable.  e) Described in 2.6. and 2.7. sections of Methods. |
| 7. **Assessment of assumptions: Describe any methods used to assess the assumptions or justify their validity.** | Our sensitivity analyses (single SNP- and leave-one-out- analyses), tests for instrument heterogeneity (Cochran’s Q test and funnel plots), horizontal pleiotropy and outlier analyses (MR-Egger tests, MRPRESSO tests, radial plots) are described in detail in sections 2.6 – 2.9 of Methods. |
| 8. **Sensitivity analyses:** Describe any sensitivity analyses or additional analyses performed. | Our test of instrument sensitivity is described in 2.3. section of Methods. Post-hoc sensitivity analyses of causal estimates are described in 2.6. and 2.7. sections of Methods. |
| 9. **Software and pre-registration**   a) Name statistical software and package(s), including version and settings used.  b) State whether the study protocol and details were pre-registered (as well as when and  where). | a) All statistical software and packages used are described in Methods section.  b) The analysis plan is described in Methods. Study protocol was not registered, as this was an analysis of publicly available, summarized GWAS data. |
| **Results** |  |
| 10. **Descriptive data**   a) Report the numbers of individuals at each stage of included studies and reasons for exclusion. Consider use of a flow-diagram.  b) Report summary statistics for phenotypic exposure(s), outcome(s) and other relevant variables (e.g. means, standard deviations, proportions).  c) If the data sources include meta-analyses of previous studies, provide the number of studies, their reported ancestry, if available, and assessments of heterogeneity across these studies. Consider using a supplementary table for each data source.  d) For two-sample Mendelian randomization:  i. Provide information on the similarity of the genetic variant-exposure associations between the exposure and outcome samples.  ii. Provide information on extent of sample overlap between the exposure and outcome data sources. | a) Information is given in 2.1. section of Methods. Other details are provided in original publications.  b) Summary statistics are available from original publications of GWASs.  c) Details are available in original publications.  d) (i) We provide this information in S2 Table: Harmonized Data.  (ii) Described in 2.2. section of Methods. |
| 11. **Main results**   a) Report the associations between genetic variant and exposure, and between genetic variant and outcome, preferably on an interpretable scale (e.g. comparing 25th and 75th percentile of allele count or genetic risk score, if individual-level data available).  b) Report causal effect estimate between exposure and outcome, and the measures of uncertainty from the MR analysis. Use an intuitive scale, such as odds ratio, or relative  risk, per standard deviation difference.  c) If relevant, consider translating estimates of relative risk into absolute risk for a meaningful time-period.  d) Consider any plots to visualize results (e.g. forest plot, scatterplot of associations between genetic variants and outcome versus between genetic variants and exposure). | a) Associations between IVs and exposures as well as IVs and outcome are presented in S2 Table: Harmonized Data.  b) Our results are given including odds ratios and confidence intervals in 3.2. section of Results and Table 2.  c) Not applicable  d) We visualize results using scatter plots in Figure 1 and S1 Figure and forest plots in Figure 2 and S2 Figure. |
| 12. **Assessment of assumptions**   a) Assess the validity of the assumptions.  b) Report any additional statistics (e.g., assessments of heterogeneity, such as I2, Q statistic). | a) We assess the validity using sensitivity analyses and results are presented in 3.4. and 3.5. sections of Results.  b) We report results from heterogeneity analyses in 3.6. section of Results. Findings from horizontal pleiotropy and outlier analyses are described in 3.7. section of Results. |
| 13. **Sensitivity and additional analyses**   a) Use sensitivity analyses to assess the robustness of the main results to violations of the assumptions.  b) Report results from other sensitivity analyses (e.g., replication study with different dataset, analyses of subgroups, validation of instrument(s), simulations, etc.).  c) Report any assessment of direction of causality (e.g., bidirectional MR).  d) When relevant, report and compare with estimates from non-MR analyses.  e) Consider any additional plots to visualize results (e.g., leave-one-out analyses). | a) Described in 3.4 – 3.7. sections of Results.  b) Validation of instruments was assessed using F-statistic and presented for each SNP in S2 Table: Harmonized Data  c) We visualized the directions of causal associations using scatter plots (Figure 1 and S1 Figure). As previous bidirectional MR studies confirmed that the causal association is unidirectional (increased BMI causes PCOS but not vice versa), we assumed this holds in the current analysis, and bidirectional analyses were not conducted.  d) We compared with findings from observational studies in Discussion.  e) Additional plots include Figure 3 – 5 and S3 Figure – S5 Figure (leave-one-out analysis plots, funnel plots, radial plots). |
| **Discussion** |  |
| 14. **Key results** | We describe key results in the first paragraph of Discussion. |
| 15. **Limitations**   Discuss limitations of the study, taking into account the validity of the MR assumptions, other sources of potential bias, and imprecision. Discuss both direction and magnitude of any potential bias, and any efforts to address them. | We discuss these aspects in Discussion: paragraph 2 |
| 16. **Interpretations**   a) Give a cautious overall interpretation of results considering objectives and limitations.  Compare with results from other relevant studies.  b) Discuss underlying biological mechanisms that could be modelled by using the genetic  variants to assess the relationship between the exposure and the outcome.  c) Discuss whether the results have clinical or policy relevance, and whether interventions  could have the same size effect. | a) Discussion: paragraphs 1 – 5.  b) Discussion: paragraphs 3 – 5.  c) Discussion: paragraph 7. |
| 17. **Generalizability:** | Discussion: paragraph 2. |
| 18. **Funding:** | Provided in Declarations section. |
| 19. **Data and data sharing:** | We used publicly available data and links have been provided. |
| 20. **Conflicts of Interest:** | Authors have declared no conflicts of interest. |
